# Accuracy of a Risk Alert Threshold for ICU Hypoglycemia: Retrospective Analysis of Alert Performance and Association with Clinical Deterioration Events

**DOI:** 10.1101/2022.06.15.22276435

**Authors:** William B. Horton, Elaine E. Hannah, Frances L. Morales, Cherie R. Chaney, Katy N. Krahn, Pavel Chernyavskiy, Matthew T. Clark, J. Randall Moorman

## Abstract

**Objective:** To quantify the accuracy of and clinical events associated with a risk alert threshold for impending hypoglycemia during ICU admissions.

**Design:** Retrospective electronic health record review of clinical events occurring ≥1 and ≤12 hours after the hypoglycemia risk alert threshold was met.

**Setting:** Adult ICU admissions from June 2020 through April 2021 at the University of Virginia Medical Center.

**Patients:** 342 critically-ill adults that were 63.5% male with median age 60.8 years, median weight 79.1 kg, and median body mass index of 27.5 kg/m^2^.

**Interventions:** Real-world testing of our validated predictive model as a clinical decision support tool for ICU hypoglycemia.

**Measurements and Main Results:** We retrospectively reviewed 350 hypothetical alerts that met inclusion criteria for analysis. The alerts correctly predicted 48 cases of Level 1 hypoglycemia that occurred ≥1 and ≤12 hours after the alert threshold was met (positive predictive value= 13.7%). Twenty-one of these 48 cases (43.8%) involved Level 2 hypoglycemia. Notably, three myocardial infarctions, one medical emergency team call, two initiations of cardiopulmonary resuscitation, 6 unplanned surgeries, 19 deaths, 20 arrhythmias, and 38 blood or urine cultures were identified or obtained ≥1 and ≤12 hours after an alert threshold was met. Alerts identified 102 total events of hypoglycemia and/or clinical deterioration, yielding a positive predictive value for any event of 29.1%.

**Conclusions:** Alerts generated by a validated ICU hypoglycemia prediction model had positive predictive value of 29.1% for hypoglycemia and other associated adverse clinical events.

**Key Points:** *Question:* What are the accuracy of and clinical events associated with a risk alert threshold for ICU hypoglycemia?

*Findings:* We retrospectively reviewed 350 hypothetical alerts that correctly predicted 48 cases of Level 1 hypoglycemia occurring ≥1 and ≤12 hours after the alert threshold was met (positive predictive value= 13.7%). Notably, three myocardial infarctions, one medical emergency team call, two initiations of cardiopulmonary resuscitation, 6 unplanned surgeries, 19 deaths, 20 arrhythmias, and 38 blood or urine cultures were identified or obtained ≥1 and ≤12 hours after an alert threshold was met.

*Meaning:* Alerts generated by a validated ICU hypoglycemia prediction model had positive predictive value of 29.1% for hypoglycemia and other associated adverse clinical events.

## Introduction

Randomized controlled trials demonstrate that intensive care unit (ICU) hypoglycemia is strongly associated with increased morbidity and mortality (1). The well-established biochemical, hemodynamic, and electrophysiological changes that occur during hypoglycemia (2) make it an ideal target for predictive analytics monitoring; however, few studies have focused on model development specifically for ICU hypoglycemia (3-5). We recently described a pathophysiologic signature of impending ICU hypoglycemia that incorporated hemodynamic and electrophysiological bedside monitoring data in a logistic regression model (6). A necessary step in translating this model to clinical practice is understanding how it would perform when operationalized as a real-time alert. Towards this goal, the aim of the current study was to retrospectively quantify the accuracy of and identify the nature of clinical events associated with large, abrupt increases (*i.e*., spikes) in hypoglycemia risk during ICU admissions at the University of Virginia (UVA) Medical Center.

## Methods

We performed a retrospective analysis of adult ICU admissions where Prediction Assistant, CoMET® *inside* (Premier, Inc.; Charlotte, NC) was in place from June 2020 through April 2021 at UVA Medical Center. Prediction Assistant collected laboratory results and flowsheet vital signs from the electronic health record (EHR) along with continuous cardiorespiratory monitoring data from the UVA Kafka System in real-time (7). Prediction Assistant used these data to estimate the relative risk of impending ICU hypoglycemia based on our validated multivariable logistic regression model containing 41 independent predictors (6). For this, the model was employed in the current cohort to estimate the probability of hypoglycemia in the next 12 hours, then that probability was divided by 0.00436 (*i.e*., the average probability of hypoglycemia in the next 12 hours). This study (“Chart Review for Predictive Modeling in the Hospital”; IRB #22152), was reviewed by the UVA IRB for Health Sciences Research and the need for IRB approval and informed consent was waived on 08/07/2020. All procedures were followed in accordance with the Helsinki Declaration of 1975.

We analyzed relative risk estimates every 15 minutes. We focused on large spikes in risk defined as an increase of ≥10 units compared with the average 2-3 hours prior (Figure 1). For example, the threshold was met if risk increased from 2 to 12 units or from 0.3 to 10.3 units. We examined only the first alert threshold met during an ICU admission (*i.e*., if multiple thresholds were met in one admission, only the first was examined and the remainder were excluded).

**Figure 1.**
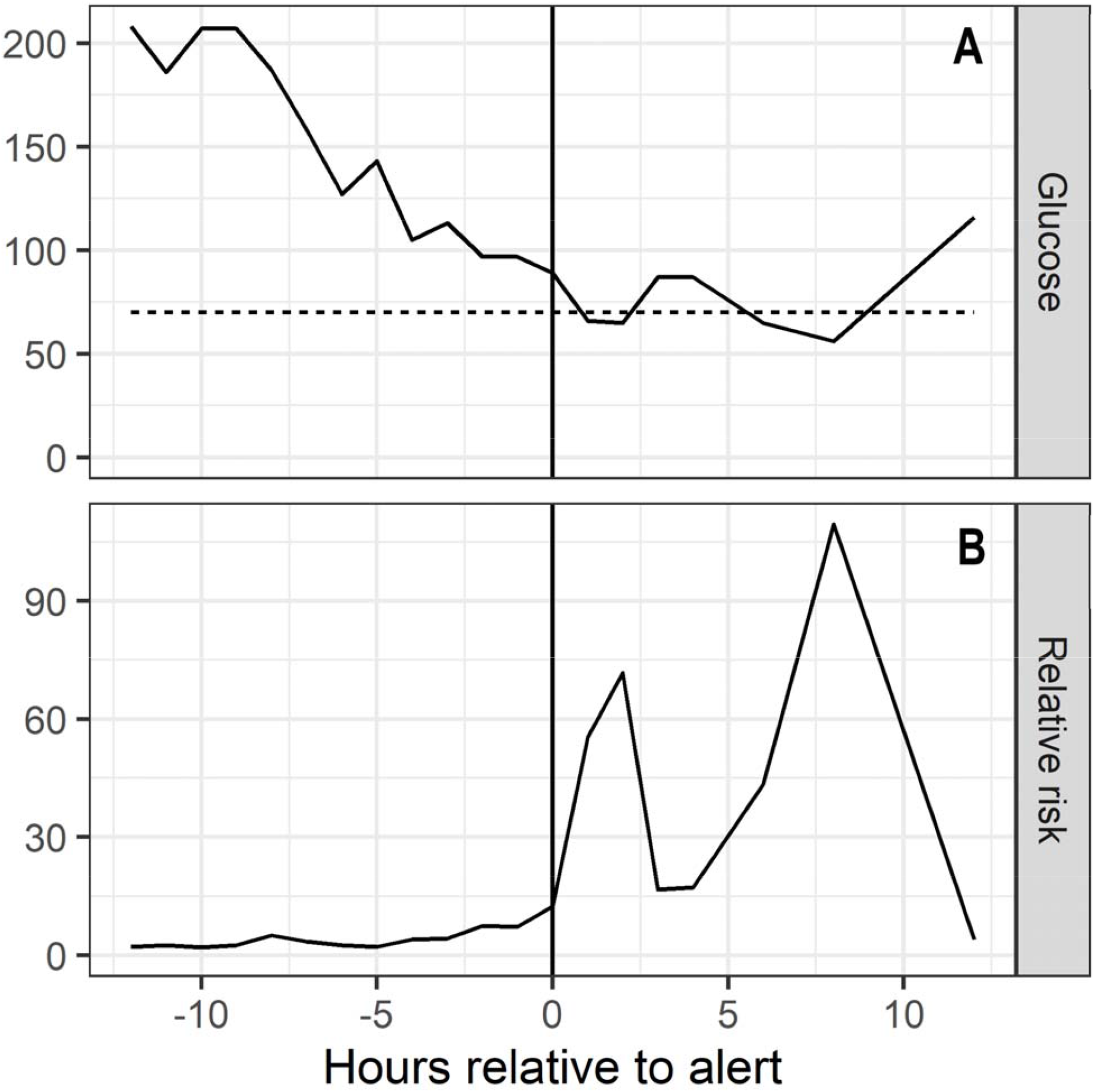
Time series demonstrating fingerstick glucose (Panel A) and hypoglycemia relative risk (Panel B) values around the time an ICU hypoglycemia alert threshold was met. Note that the alert threshold (solid vertical line in Panels A and B) was met prior to onset of Level 1 hypoglycemia (*i.e*., <70 mg/dL; represented by the dashed horizontal line in Panel A).

We also examined the EHR for clinical events associated with these risk spikes. For this, we assessed the period of time ≥1 but ≤12 hours after the spike. We considered this enough time to allow clinicians to see patients and intervene (*i.e*., no less than one hour) but not so long as to lose association of the spike with the event (*i.e*., more than 12 hours). Hypoglycemia categories were consistent with those recommended by the American Diabetes Association’s *Standards of Medical Care in Diabetes* (8): Level 1 hypoglycemia was a blood or fingerstick glucose value <70 mg/dL (3.9 mmol/L) and ≥54 mg/dL (3.0 mmol/L), and Level 2 hypoglycemia was a blood or fingerstick glucose value <54 mg/dL (3.0 mmol/L). We recorded the time of first Level 1 and/or Level 2 hypoglycemia as well as whether intravenous dextrose, oral glucose tablets, and/or liquid sugar (*e.g*., orange juice) were administered as treatment and whether the subcutaneous insulin dose was adjusted. We also noted other clinical deterioration events: medical emergency team call/visit, myocardial infarction, initiation of cardiopulmonary resuscitation, unplanned surgery, death, obtaining a blood or urine culture, and/or arrhythmia on electrocardiogram. Statistical analyses were performed in GraphPad Prism (GraphPad Software, San Diego, California).

## Results

### Identification and Treatment of ICU Hypoglycemia

Table 1 provides demographic and clinical data for the study cohort (n=342 patients). Notably, the cohort was 63% male, had median age of ∼61 years, and was distributed amongst 5 different ICUs. The median increase in hypoglycemia relative risk prior to an alert was 10.57.

**Table 1.** Demographic and clinical characteristics of the study cohort (n=342). (CCU= coronary care unit; MICU= medical intensive care unit; CICU= cardiovascular intensive care unit; STICU= surgical-trauma intensive care unit; TCVPO= thoracic cardiovascular post-operative unit; IQR= interquartile range).

| Variable | Value |
| --- | --- |
| Sex, *n* (%) |  |
| Male | 217 (63.45) |
| Female | 125 (36.55) |
| Race, *n* (%) |  |
| White | 223 (65.20) |
| African-American | 90 (26.32) |
| Other | 19 (5.56) |
| Unspecified | 5 (1.46) |
| Asian | 3 (0.88) |
| Multi-Race | 2 (0.58) |
| Intensive Care Unit, *n* (%) |  |
| CCU | 85 (24.29) |
| MICU | 64 (18.29) |
| CICU | 66 (18.86) |
| STICU | 87 (24.86) |
| TCVPO | 48 (13.71) |
| Age, median years (IQR) | 60.82 (49.94-71.88) |
| Weight, median kilograms (IQR) | 79.10 (66.25-96.75) |
| BMI, median kg/m<sup>2</sup> (IQR) | 27.47 (23.64-32.83) |
| Rise In Hypoglycemia Relative Risk, median (IQR) | 10.57 (10.24-11.33) |

We reviewed 589 total threshold alerts, 350 (59.4%) of which were first alerts for the ICU admission. These alerts correctly predicted 48 cases of Level 1 hypoglycemia (positive predictive value [PPV]=13.7%), 21 (44%) of which involved Level 2 hypoglycemia. During the study period, there were 199 hypoglycemic episodes for which no alert was dispatched (sensitivity= 19.4%). We note here, however, that for this study the hypothetical alert system was calibrated to dispatch only two alerts per day which limited overall sensitivity.

All 48 hypoglycemia cases were treated with intravenous dextrose. Three (6.3%) were treated with liquid sugar via oral or enteral access, while none were treated with oral glucose tablet administration. In seven cases (14.6%), the subcutaneous insulin regimen was adjusted <12 hours after the alert.

### Association of Clinical Deterioration Events with ICU Hypoglycemia Alerts

We identified 3 myocardial infarctions, one medical emergency team call, two initiations of cardiopulmonary resuscitation, 6 unplanned surgeries, 19 deaths, 20 arrhythmias, and 38 blood or urine cultures following an alert. Overall, the predictive analytics monitoring alert identified 102 patients with hypoglycemia, clinical deterioration, or both, resulting in a 29.1% PPV for any event.

## Discussion

Appropriate glycemic control is a necessary but often overlooked component of quality-driven inpatient healthcare. However, ICU hypoglycemia is consistently linked to greater morbidity and mortality and is routinely identified as the limiting factor for improving glycemic control (1, 9). These points emphasize the need for a more proactive approach to ICU hypoglycemia. Here, we studied a predictive model’s ability to prospectively identify both hypoglycemic and clinical deterioration events in ICU patients and found that our risk alert threshold had a PPV of 13.7% for true hypoglycemia events. The PPV performance of our alert threshold is in-line with similar studies in this field. For example, Mathioudakis et al. developed and validated a machine learning model to predict near-term hypoglycemia risk in non-ICU patients (10) that achieved PPV values of 9% during internal validation and 12-13% during external validation. Our alert threshold achieved a similar PPV for hypoglycemia in an ICU population, providing important data as we seek to incorporate predictive analytics monitoring into clinical trials of a prospective alert system.

We also identified numerous adverse clinical events that occurred after an alert, including cardiac arrhythmia, myocardial infarction, cardiopulmonary resuscitation, unplanned surgery, and death. While the current study was not designed to assess whether hypoglycemia directly caused these adverse clinical events, there are numerous reports that support their relationship. For example, the pronounced sympathoadrenal activation during hypoglycemia is known to cause abnormal cardiac repolarization that can induce cardiac arrhythmias (2).

Our study has several limitations that should be noted. First, data collection was limited to one tertiary academic medical center with a high proportion of medically-complex patients that may limit generalizability. Second, we note that risk spikes of ≥10 have different meanings depending on the baseline level. For example, a rise from 0.1 to 10.1 is a larger relative increase in probability than a rise from 2 to 12. Nonetheless, we felt this was a clinically-acceptable approach for future trials and implementations. Finally, we developed and validated our predictive model in insulin-treated ICU patients but the alert threshold was employed in all ICU patients for the current study.

## Conclusion

Alerts generated by our validated ICU hypoglycemia prediction model had positive predictive value of 29.1% for hypoglycemia and other associated adverse clinical events. To complete impact analysis, we are planning a cluster-randomized controlled clinical trial where we will incorporate our model into a prospective alert system and test its impact on hypoglycemia and associated endpoints like mortality and length-of-stay.

## Data Availability

All data produced in the present study are available upon reasonable request to the authors

